# Personalized oral care (Precaries): an intervention study customized according to genetic cause and risk

**DOI:** 10.1101/2024.01.23.24300787

**Authors:** Anna Westerlund, Halah Khalifa, Ranna Yousif, Gustavo Araujo, Evelina Lundqvist, Erica Larsson, Rakel Thrastardottir, Rebecka Akhlaghi, Victoria Granciuc, Charlotta Svanberg, Maria André, Anna Lehrkinder, Farhan Bazargani, Christina Goriel Radsjö, Caspar Carlfjord, Anke Krämer, Niels Ganzer, Isabell Hansson, Erik Frilund, Eva Josefsson, Rune Lindsten, Anders Magnusson, Brygida Grunwald, Firas Hittini, Nurije Kryeziu, Henning Looström, Mikael Sonesson, Reem Al-Taha, Hanna Surac, Seifi Esmaili, Haris Isic, Anna Tegnell, Samuel Andersson, Mai Lin Lövgren, Jenny Kallunki, Anna E Lorenzo, Elena Arezzo, Agata Jasna, Tumkur Sitaram Raviprakash, Eva Strömqvist-Engbo, Amanda Burstedt, William Rosenbaum, Oscar Öhman, Wolfgang Lohr, Ann-Charlotte Rönn, Tomas Axelsson, Lena Mårell, Lennart Österman, Edward J Hollox, Ulrika Westerlind, Karina Persson, Henrik Clausen, Per Liv, Patrik Rydén, Johan Henriksson, Laura Carroll, Nongfei Sheng, Pär Larsson, Nicklas Strömberg

**Author notes:** Second authors that have contributed equally (Halah Khalifa, Ranna Yousif, Gustavo Araujo, Evelina Lundqvist, Erica Larsson, Rakel Thrastardottir).

## Abstract

**Introduction:** Dental caries is a disease that affects billions of people, and involves high and low genetic susceptibility phenotypes and different causal subtypes. The randomized clinical trial Precaries-RCT will evaluate caries prevention in adolescents, customized according to genetic cause and risk. Here we describe the Precaries-RCT and two nested Precaries studies for cost-efficient oral healthcare and personalized dentistry.

**Methods and analysis:** Here we present a basic and adaptive protocol for the Precaries-RCT multicentre caries intervention study, customized according to genetic cause and risk. It includes prescreening for high versus low genetic caries susceptibility, through self-performed sampling by mail of up to 2000 adolescents aimed for orthodontic treatment at community clinics, of which 520 are enrolled in the RCT. The participants are allocated into two groups – a high and a low genetic caries susceptibility group – that each is assigned to intensive or standard prevention. The primary outcome is % reduction in caries increment, relative to prevention and genotype, with caries outcomes measured using tactile and visual methods, bitewing radiographs, clinical photos, and quantitative laser fluorescence. The adaptive design allows for determination of incidence and progression rates and for inclusion of additional human and microbiota biomarkers and study subjects. Biological samples (e.g. swab DNA, whole and parotid saliva, and microbiota) and questionnaire data are collected. Here we also outline the nested Precaries-adolescent sample for mining of predictor and therapeutic target genes and Precaries-birth cohort samples for implementation of our findings in childhood.

**Ethics and dissemination:** Ethical approval was obtained from the Swedish national board research ethics committee (Dnr 2020-02533). Informed consent will be obtained from each participant. The findings will be disseminated to the public through conference presentations and publication in peer-reviewed scientific journals.

**Trial registration number:** www.clinicaltrials.gov, NCT05600517

**STRENGTHS AND LIMITATIONS OF THIS STUDY:**

- Caries classification and prevention customized according to genetic cause and risk, and caries outcome measurements by tactile and visual methods, bitewing radiographs, clinical photos, and quantitative laser fluorescence.
- Multicentre study with orthodontic patients and conditions representative of the Public Dental Service clinics in Sweden, facilitating implementation, though in a part of the population.
- Prospective study design in an orthodontic model with shortened study time and caries development on available smooth tooth surfaces. Frequent follow-up enables study termination of individuals with high caries progression, which may allow further shortening of study times, but with reduced individual data for the entire study period.
- Consensus-based intensified and self-care prevention in multiple repeated blocks, ensuring a high therapeutic dose; and basic and adaptive design, allowing flexible study time, caries incidence and progression outcomes, and extensive genetic profiling for online multimodal machine learning.
- Synergizes with the Precaries-adolescence sample for mining of caries predictors and therapeutics, and Precaries-birth cohort for implementation in primary dentition/childhood.

## INTRODUCTION

In populations with a high prevalence, dental caries is considered a lifestyle disease, related to poor diet and oral hygiene [1, 2]. However, in Sweden and most Western countries, preventative treatment and other advances have transformed caries-afflicted populations to healthy populations with physiological (lifestyle) homogeneity but genetic heterogeneity [3–5]. In low-prevalence populations, 85% of individuals are free of caries or exhibit low-to-moderate levels, and may thus represent subjects with low genetic susceptibility and lifestyle-dependent caries [5–7]. The remaining 15% are high-caries individuals with recurrent caries and that do not respond to prevention, who may represent subjects with high genetic susceptibility and the immunodeficiency type of caries [6, 7]. Personalized dentistry will customize risk assessment, prevention, and treatment according to the individual’s genetic and lifestyle profile.

Revisiting causes of dental caries in the Swedish population, and other populations with a low caries prevalence, may help identify caries-associated genes and pathways for prediction and therapeutic approaches [6, 7]. We have identified saliva immunodeficiency and lifestyle causal subtypes of caries in persons with phenotypes of high genetic susceptibility (P4a^+^) and low genetic susceptibility (P4a^−^), respectively, based on variations in *PRH1* and *PRH2*, within a prospective sample of 452 adolescents [6] (figure 1). Highly susceptible P4a^+^ phenotypes with immunodeficiency-type caries likely account for a portion of the high-caries population. Immunodeficiency-type caries phenotype may differ both in adhesion and barrier functions, and sensitivity to pioneer microbiota on teeth [6, 7]. DMBT1 is another self versus non-self pattern recognition molecule that is associated with caries [8, 9]. In the sample of adolescents, we also identified a *Streptococcus mutans* commensal pathogen-dependent subtype of caries that occurs in persons with a certain host-microbe adhesin-receptor genetic variation [7]. The low- and high-caries phenotypes are stable trajectories of a plausible genetic nature in the primary and permanent dentitions [10]. Therefore, targeting the underlying human and oral microbiota genes and polymorphisms may facilitate the development of personalized dentistry and oral care.

**Figure 1.**
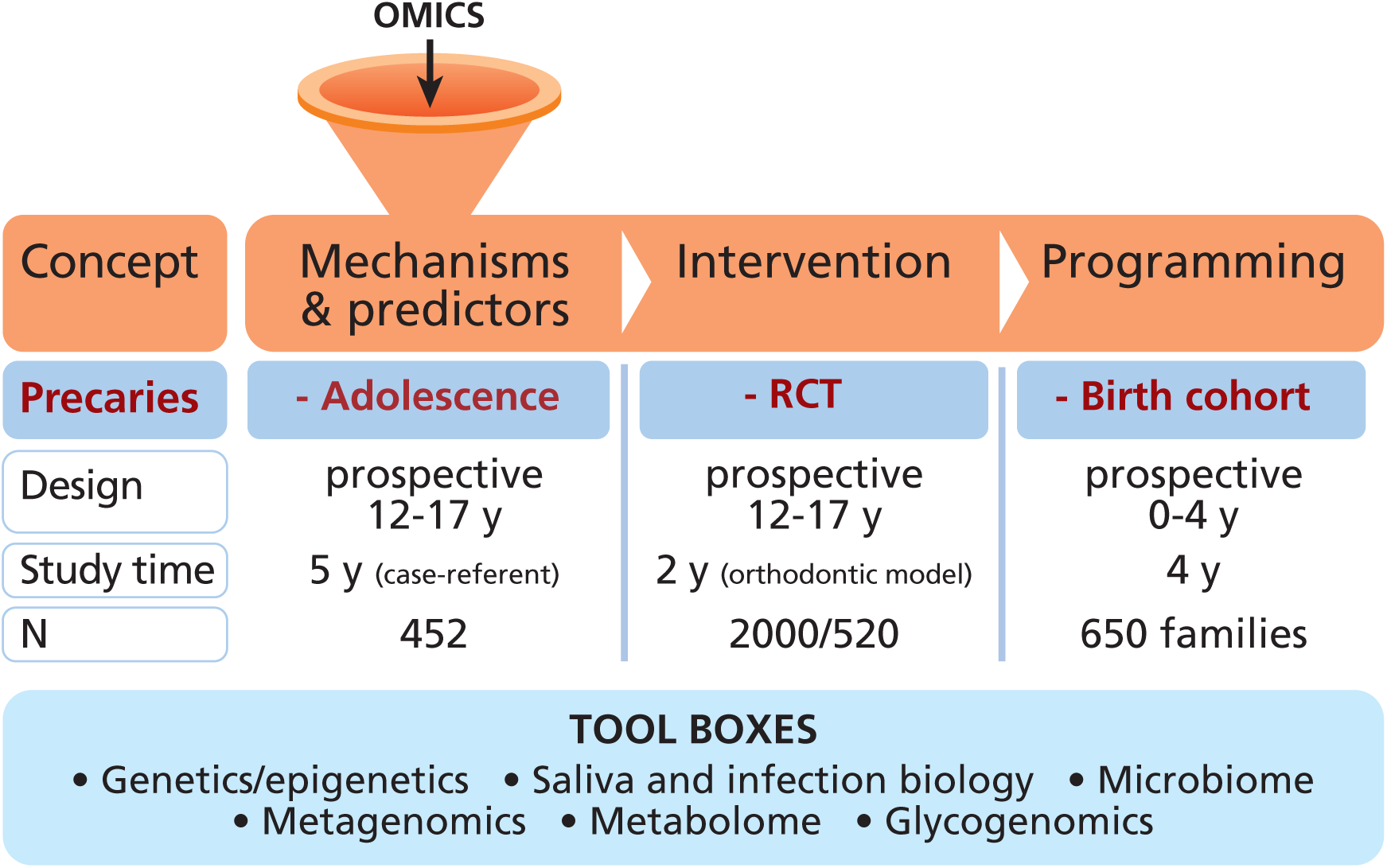
The Precaries-RCT and nested study samples. The prospective case-referent Precaries-adolescence sample is used for mining of predictor and therapeutic target genes. The basic and adaptive Precaries-RCT uses an orthodontic caries model with shortened study time, and is used to identify predictors and responsiveness to prevention. In the Precaries-birth cohort, predictors and responsiveness are evaluated and implemented in the primary dentition.

Risk prediction is performed to classify individuals into sick and healthy groups for cost-effective oral care management, or to identify a person’s fine-tuned individual risk to provide personalized prevention and treatment. However, there is a lack of tools for the classification and prediction of individual risk, before caries lesions develop and become irreversible [11, 12]. Early risk prediction may contribute to overcoming the non-responsiveness of individuals with recurrent caries to standard fluoride prevention, which prevents only 15% of caries progression [3, 4]. Notably, fluoride is on the list of neurotoxic agents that may impair neurophysiological development in childhood [13].

There are large knowledge gaps regarding the causal subtypes, prediction, and prevention of caries [12, 14]. However, rapidly developing life science technologies enable targeted approaches to identifying caries- and health-associated genes and molecular pathways that may be useful as predictors and therapeutic agents, to facilitate the development of personalized or precision dentistry [15–23]. For example, the *PRH1* and *PRH2* predictor alleles encode acidic proline-rich proteins and prebiotic peptides [6, 24]. Precision dentistry is also facilitated by untargeted omic approaches using all biological and clinical data in deep learning and AI methods [21, 22]. We can also foresee the development of novel ways to control the microbiota at infancy and childhood, when immunity matures in response to genetic, exposome, and epigenetic factors [25]. The identification of human and microbiota target genes and pathways for causal subtypes will lead to the generation of both predictors and therapeutics.

One limitation in studies on caries causes, predictors, and therapeutics is the slow progression of caries (3-5 years), and the lack of methods to measure caries induction and progression. In this respect, treatment with orthodontic multibrackets—which accumulate bacteria and disrupt the anterior-to-posterior flow of saliva immunity in oral biofilms— coincides with rapid caries development (6-12 months) on available, unaffected, smooth tooth surfaces [26–29]. However, orthodontic treatment coincides with rapid caries in genetic susceptible adolescents particularly, while those with low genetic susceptibility develop less caries due to the comparably high prevention given during the treatment [6,7]. Thus, the orthodontic caries model will shorten study time, and facilitate studies on caries induction and progression.

This protocol describes the Precaries-RCT study for evaluation and implementation of caries prevention customized according to genetic cause and risk (figures 1 and 2). The study is a multicentre, randomized, two-arm, controlled trial of intensified and self-care prevention of caries among Swedish adolescents, with an adaptive design. The protocol also describes the nested Precaries-adolescence sample for mining of human and microbiota target genes, and the Precaries-birth cohort study for implementation in childhood.

### Aims and hypothesis

The overall purpose of the Precaries-RCT study is to evaluate caries prevention customized according to genetic cause and risk, and initiated before clinical symptoms. The primary hypotheses are that intensified prevention must be started before clinical symptoms to prevent caries development in high-caries phenotypes with high genetic susceptibility, and that traditional standard prevention is sufficient to prevent caries development in low-to-moderate-caries phenotypes with low genetic susceptibility. Correspondingly, the primary aims are to show that intensified prevention started before clinical symptoms emerge can prevent caries development in high-caries P4a^+^ phenotypes with high genetic susceptibility, and to demonstrate that traditional standard prevention is sufficient to prevent caries in low-to-moderate-caries P4a^−^ phenotypes with low genetic susceptibility. The secondary aims are as follows: to show that *PRH1* and *PRH2* variations predict high-caries versus low-caries phenotypes, that adaptive inclusion of additional human and microbiota genotypes can improve risk assessment and responsiveness to prevention, that improved caries induction and progression measures can be developed, and that inflammatory responsiveness and bone loss can be used to identify potential predictors of adult periodontal disease, as well as to determine the frequency of mineralization disturbances.

## METHODS AND ANALYSIS

### Study design and setting

Precaries-RCT is a multicentre, randomized, controlled, two-arm parallel, and adaptive clinical intervention trial with an allocation ratio of 1:3 (figure 2, supplemental table S1), based on the nested Precaries-adolescence study [6,7] (figure 1, supplemental table S2) Participants in Precaries-RCT are enrolled from about 10 specialist clinics of orthodontics from the Public Dental Services in several regions (counties) throughout Sweden (figure 3 and online supplement, section 3). The intervention phase is up to 2 years long. Supplemental table 1 presents the summary schedule for enrolment, interventions, and assessments; and figure 4 and supplemental figure S1 presents flowcharts of the study participants and intervention repeat periods.

**Figure 2.**
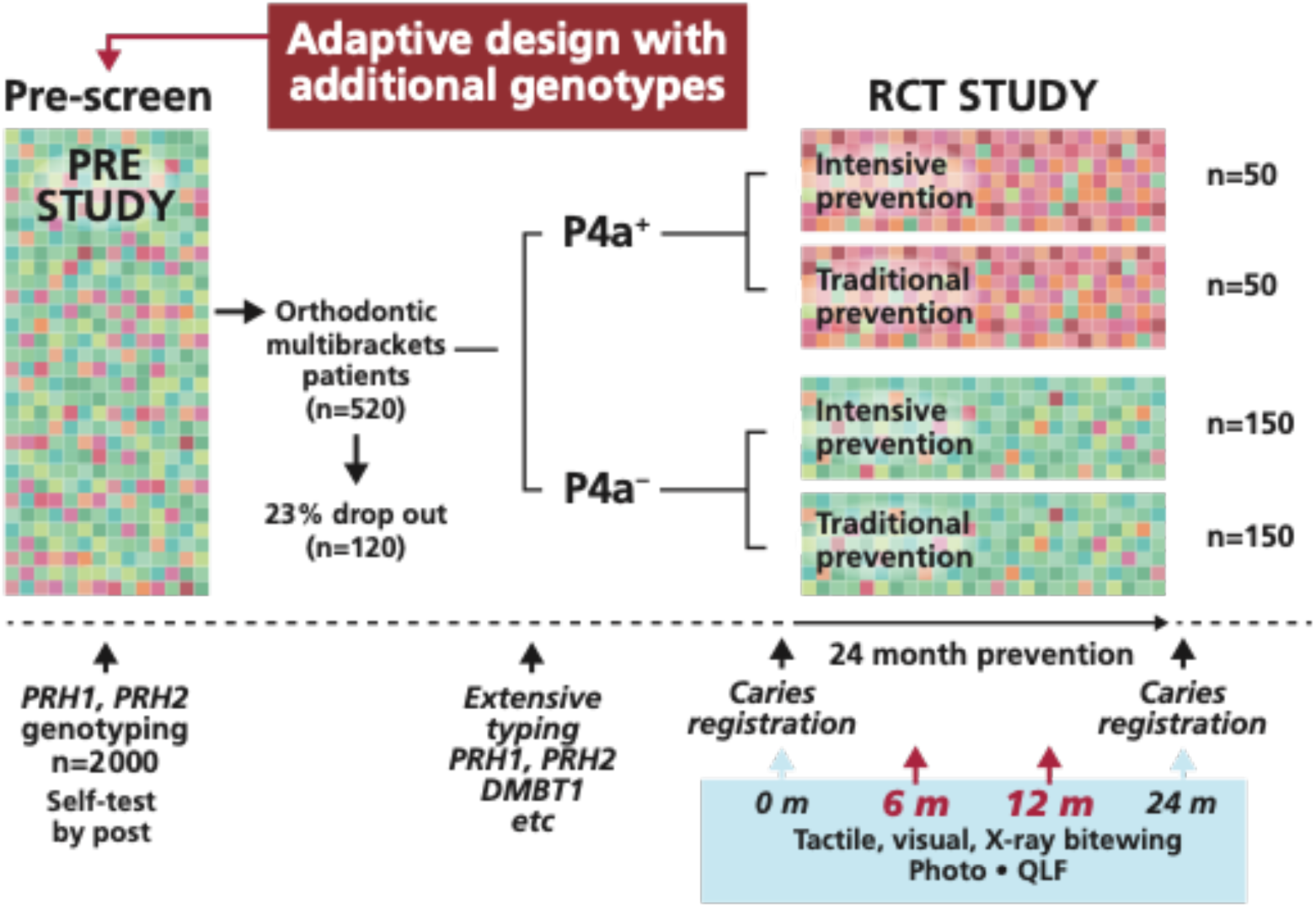
The Precaries-RCT study. The basic RCT design includes a prescreening component, and a RCT part with high genetic susceptibility (P4a^+^) and low genetic susceptibility (P4a^−^) groups that each receive intensified or traditional prevention during 24 months, with measurements at 0 and 24 months. The adaptive design allows registration of outcomes at 6 and 12 months for a flexible study time, and for determination of caries incidence and progression rates. The adaptive design also allows the evaluation of additional predictor target genes, and inclusion of additional study subjects to meet power needs.

**Figure 3.**
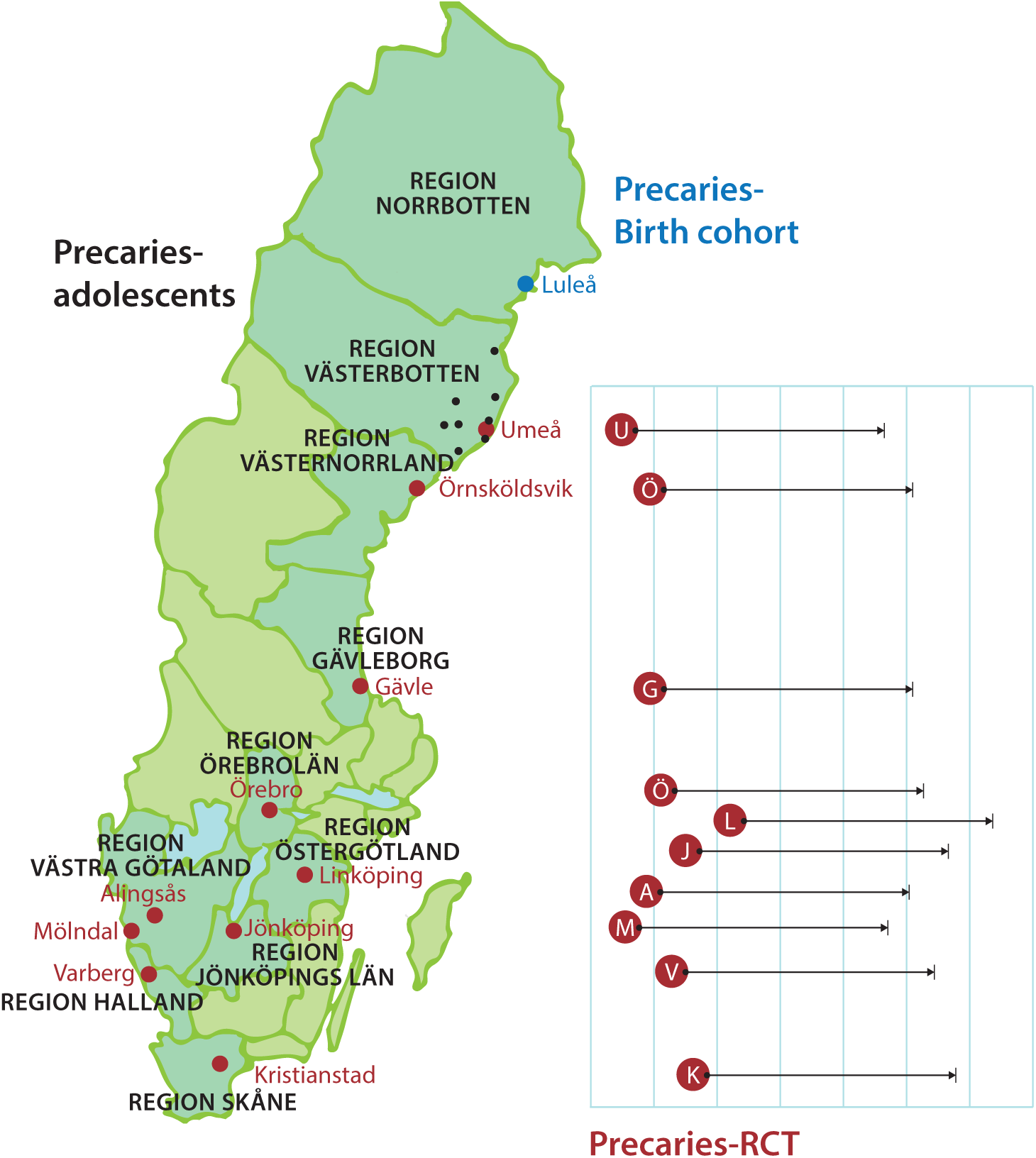
Ten clinical sites (red dots) and eight regions (counties) participating in the Precaries-RCT study. The relative start-up and time scheduling of the clinics are presented on the right. Region Västerbotten is the site for the Precaries-adolescents cohort, sampled at clinics in Umeå and in the region (black dots). Also marked are the region and clinical site (blue, Luleå) for the Precaries-birth cohort.

**Figure 4.**
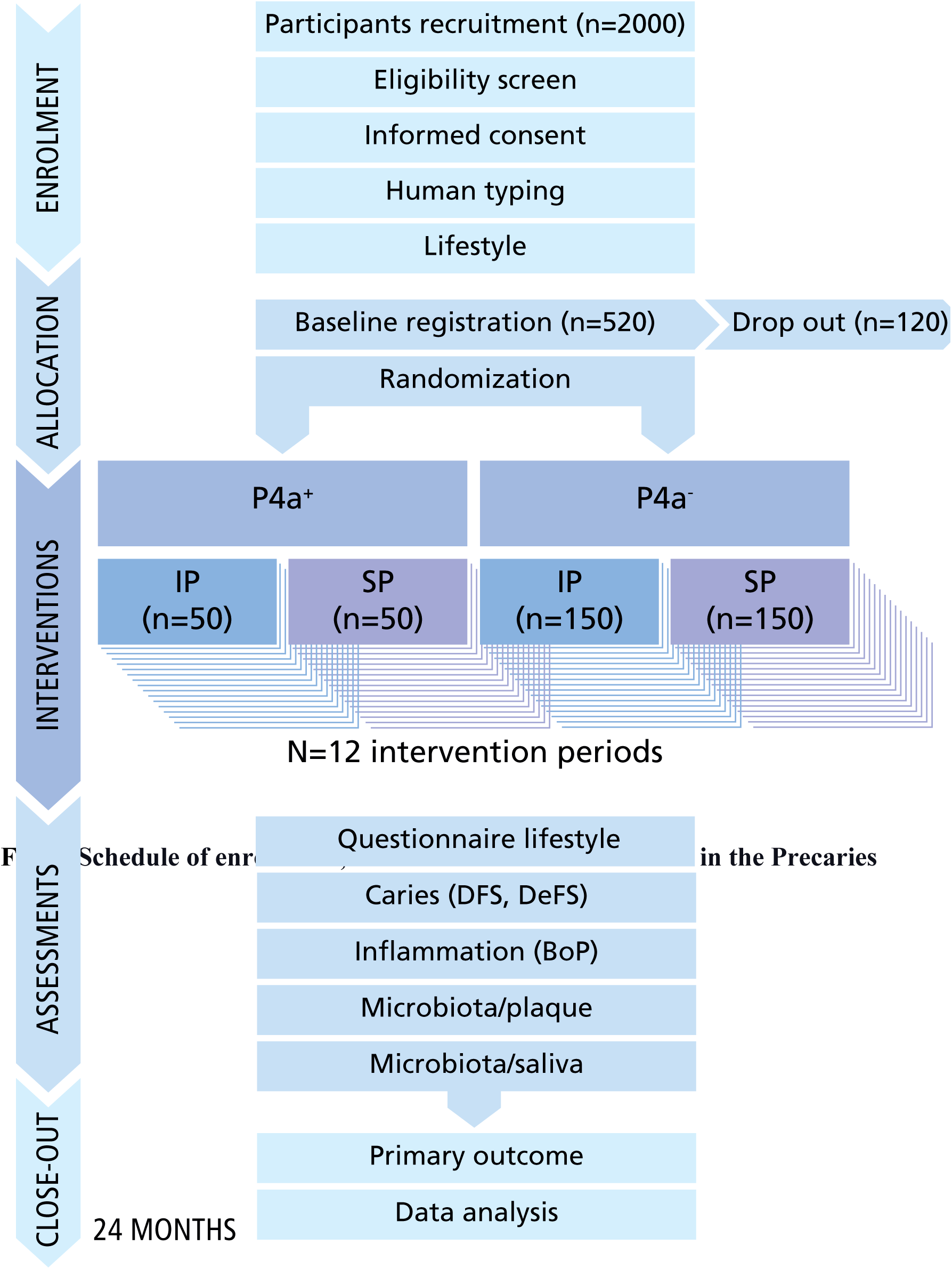
Flowchart of enrolment, allocation, treatment, and follow-up of study participants in the Precaries-RCT. IP=Intensive prevention; SP=Standard prevention. Interventions are provided in 12 repeated periods of 8 weeks each. Each intervention period starts with instruction and ends with a follow-up.

The Precaries-RCT comprises a prescreening of up to 2000 adolescents who are planned to start ordinary orthodontic treatment with multibrackets at specialist clinics of orthodontics, to select individuals with high genetic susceptibly (P4a^+^) and low genetic susceptibility (P4a^−^) genotypes (figures 2 and 3). The intervention portion of the RCT includes 520 adolescents who are classified into two risk groups—high genetic susceptibility to caries (P4a^+^) and low genetic susceptibility to caries (P4a^−^). Individuals in each groups are assigned to receive either intensive or traditional standard prevention (figures 2 and supplemental figure S1). The clinics are enrolled and started stepwise (figure 3), as an adjustment to the COVID/Coronavirus pandemic.

The study is steered by the PI and steering committee. The clinical portions are managed by two clinical coordinators. One coordinator manages the patient recruitment, patient and clinical data sampling (REDCap), and regular digital and physical follow-up with the clinics. The second coordinator manages the caries teams, caries calibration, and the transport of materials and clinical samples to and from clinics. At each clinic, there is a responsible dentist (typically PhD and formal specialist) and a dentist with scientific education as part of his/her specialist education. The *in* and *ex house* laboratory components of the study—including sample handling and storage, as well as statistical analyses—are managed by a laboratory and analyses steering group.

### PICO

P_atients_ Orthodontic patients with high susceptibility and low susceptibility genotypes

I_ntervention_ Fixed orthodontic appliance and intensified prevention

C_ontrol_ Fixed orthodontic appliance and standard prevention

O_utcome_ ι1DeFS, and % reduction of ι1DeFS

### Eligibility criteria

Inclusion criteria for this trial are patients 12-20 years of age, with planned orthodontic treatment using fixed multibracket appliances in the upper and lower arches. Exclusion criteria are patients with impacted canines or agenesis in the frontal region, and patients requiring maxillofacial surgery or prosthetics, due to the long treatment times.

### Recruitment and prescreening by mail

Patients are recruited at the individual specialist clinics of orthodontics (figure 3 and online supplement, section 3). Up to 2000 adolescents will be invited and prescreened using questionnaire data, as well as for P4a^+^ and P4a^−^ phenotypes by *PRH1* and *PRH2* genotyping, to meet the Precaries-RCT basic study design requirement of 520 adolescents (figure 2). Prescreening is preformed via self-performed sampling at home with buccal swabs provided by mail, together with an instructional brochure and YouTube instructional video (online supplement, section 5), and links to an online questionnaire (online supplement, section 6). The swabs are returned to our *in-house* laboratory by mail. Both the swabs and questionnaire can be complemented at the baseline visit, if needed. The prescreening enables collection of sufficient numbers of P4a^+^ adolescents, who are of lower prevalence, and adaptive inclusion of additional genotypes and subjects. It also allows estimates of genotype frequency and caries association using proxy measures (e.g. extra fluoride prevention [6]) or dental records.

### Intervention

The interventions are designed based on consensus with the participating clinics, to generate a prevention regime suitable for this study as well as for implementation in a general clinical setting (supplemental figure S1). Prevention is frequent and structured into 12 prevention periods, each 8 weeks long (figures 4 and supplemental figure S1). For all participants during the study period, toothpastes and toothbrushes are provided and renewed for free (online supplement, section 7).

Patients receiving intensive prevention receive information about self-care prevention— including diet, oral hygiene, and daily use of fluoridated toothpaste (5000 ppm)—at the start of and during treatment. The intensive prevention protocol also includes check-ups every 8 weeks, and application of topical fluoride (varnish, 22 400 ppm) at six check-ups at the clinic.

Patients receiving standard prevention receive information about self-care prevention—including diet, oral hygiene, and daily use of fluoridated toothpaste (1450 ppm)—at the start of treatment. The standard prevention protocol also includes check-ups every 8 weeks but without topical fluoride application.

### Compliance

The ordinary check-up frequency during orthodontic treatment is every 8 weeks. We expect high compliance, since children and parents are usually highly motivated. Compliance will be measured in terms of the number and percentage of fulfilled checkups, as well as based on answers to the questionnaire at baseline, 6 months, and 24 months in terms of preventive measures (online supplement, section 8).

### Randomization and blinding

Randomization will be performed using the web-based REDCap clinical data collection tool, with stratification by P4a^+^ and P4a^−^ genotype in permuted blocks based on patients per clinic. Blinding will be performed such that the dentist/nurse and patient do not know the patient’s genotype or risk group. The dentist and nurse providing the orthodontic treatment or prevention to the patient will be blinded to caries measurements by clinical photos and quantitative laser fluorescence (QLF).

### Caries registration

A caries registration team of selected dentists, including a master reference and each dentist at the individual clinics, will be trained and calibrated for the registration of DeFS (decayed, enamel included, filled surfaces in the permanent dentition) using visual-tactile registration with a mirror, probe, and four bitewing radiographs (online supplement, sections 9 and 10). At the same time-points, enamel, DeS, and lesions will also be measured using clinical photographs and quantitative light-induced QLF, as adjusted by the master reference. The dentists at the clinics will be blinded from these measurements. Examinations will be performed at baseline, 6 months, and 24 months (online supplement, sections 11-13).

The DeFS and DeS scores will be measured at the start (baseline), and at 6, 12, and 24 months, depending on the basic or adaptive study design. The increment, ΔDeFS and ΔDeS, will be calculated for each individual, relative to the unit of time. The intervention effect will be expressed as the % relative reduction in ΔDeFS caries increment. Caries induction and progression will be deduced from the rate and degree of caries at the different time-points (figure 5).

**Figure 5.**
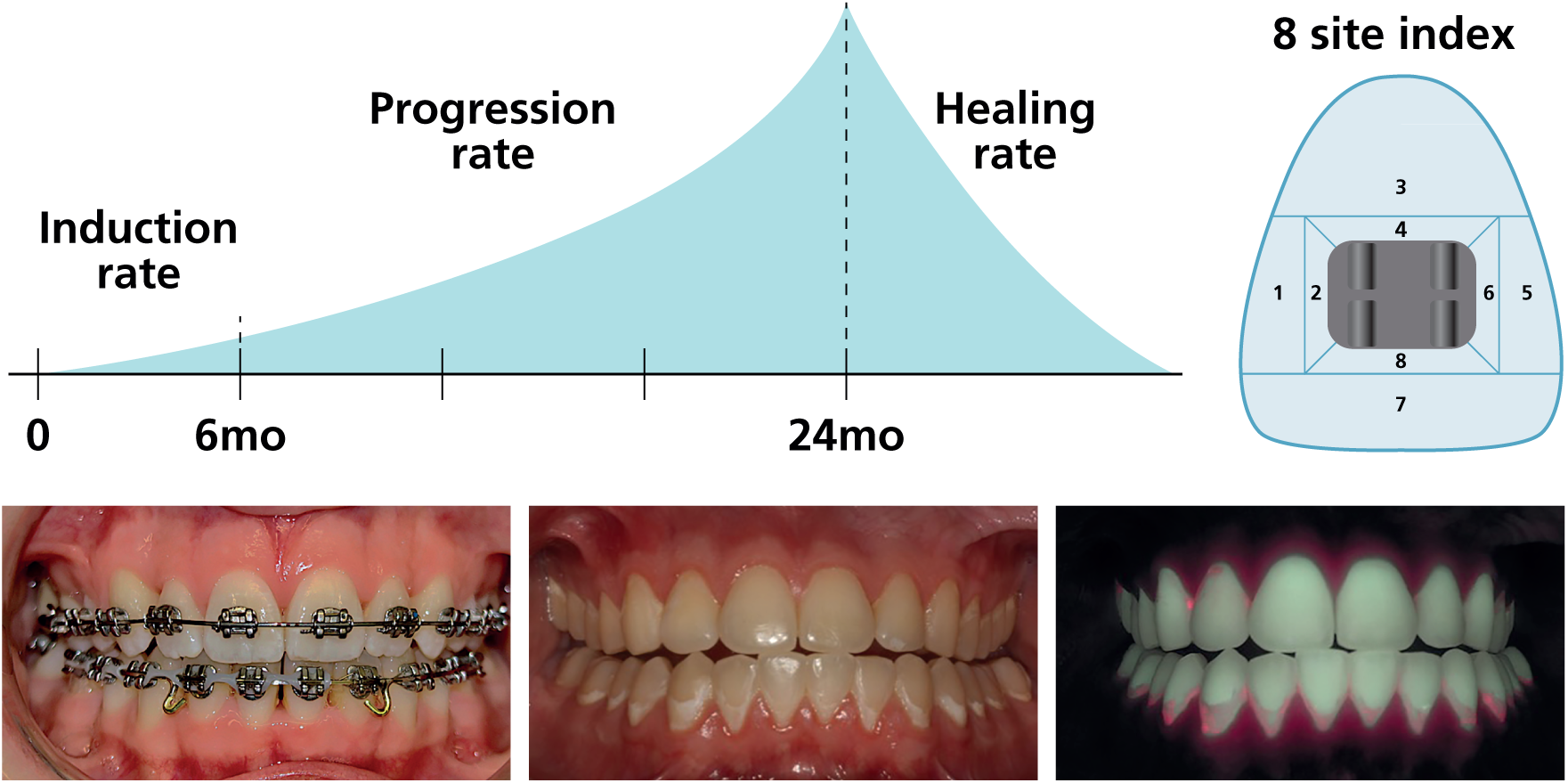
The orthodontic caries model. Schematic outline of caries outcome measures, including the rates of induction, progression, and healing (upper left), as well as an 8-site caries lesion index (upper right). The orthodontic multibrackets (lower left) accumulate bacteria and disrupt the anterior-to-posterior flow of saliva in the biofilm, which increase the risk for caries. The presence of caries is measured using tactile and visual methods, bitewing radiographs, clinical photos (middle lower), or quantitative laser fluorescence (QLF) (lower right). Treatment is terminated in cases with markedly high induction and progression of caries despite of the frequent prevention administered during the treatment period.

### Outcome variables and measures

The primary outcome variable is the % reduction in ΔDeFS caries increment over a follow-up period of 24 months relative to prevention. Secondary outcome variables are the prospective caries incidence and progression of DeS scores and rates relative to risk group, the gingival inflammation and bone loss at the end of the study, and mineralization disturbances.

For the registration of gingival inflammation and pocket depth, inflammation is measured according to bleeding on probing (BoP) at the gingival mucosa at buccal tooth surfaces in the third quadrant of the mouth. Three sites (mesial, lateral, and distal) are measured for each tooth 31-37 at time-points of 0, 2, 6, and 24 months. Bone loss is measured on X-ray radiographs at time-points of 0 and 24 months.

Disturbances in enamel mineralization are registered upon clinical examinations and in photographs and QLF. Such disturbances will include fluorosis and unclassified disturbances, and rarely (if at all) low-prevalence monogenetic diseases, such as molar-incisor-hypo-mineralization or amelogenesis imperfecta.

### Predictor variables and genotyping

The genetic predictor variables, P4a^+^ and P4a^−^ genotypes, will be measured by *PRH1* and *PRH2* genotyping using a combination of TaqMan and iPLEX MassARRAY genotyping of DNA prepared from buccal swabs collected at prescreening [6]. The Taqman, iPLEX, and originally used Illumina [6] arrays generate data with high concordance. All participants, including adolescents without calls or with some mis-match between typing methods, will be retyped by iPLEX using DNA prepared from buccal swabs collected from all patients by the dentists at the clinics at the start of the study.

### Power and sample size

The required sample size was determined using Monte Carlo simulations, assuming that ΔDeFS follows a Poisson probability distribution. Previous findings have shown a mean ΔDeFS of 5.5 for P4a^+^ children [6]. Assuming a mean ΔDeFS of 5 (to avoid undersizing), simulations indicated that 50 children in each group will be sufficient to detect a ΔDeFS reduction of 25% within the P4a^+^ arm, with 80% power, when using a Mann‒Whitney U test at a significance level of 0.05. Corresponding simulations for the P4a^−^ children revealed that 150 children in each group will be sufficient to detect a reduction of 18%, assuming a mean ΔDeFS of 2.7 [6]. Considering a dropout rate of 23%, we intend to include 130 P4a^+^ adolescents and 390 P4a^−^ adolescents.

We expect to observe a reduction of 80% or more in the ΔDeFS caries increment with an intensive prevention regimen, relative to traditional standard prevention, in both the P4a^+^ and P4a^−^ groups (figure 2).

### Adaptive design

The Precaries-RCT study has a basic (VR-KBF granting) and adaptive design (figure 2 and supplemental table S2) [30]. The adaptive design uses flexible time-points (0, 6, 12, and 24 months), and allows interim analyses of clinical outcomes, flexible study termination as study goals are reached, and the use of rate measures for DeS induction and progression. The adaptive design also allows inclusion and evaluation of further patients and predictors generated in the Precaries–adolescence sample for sick and healthy risk assessement by targeted and untargeted omics approaches (figures 2 and 6, and supplemental table S2). The prescreening facilitates recruitment of additional patients to achieve satisfactory study power and training versus test samples for on-line multimodal machine learning approaches. The adaptive design also enables the registration of other secondary clinical outcomes, such as inflammation and periodontal pockets.

### Biological sample collection and storage

At the baseline examination, prior to orthodontic appliances, a minimum of 3 mL each of stimulated parotid and whole saliva are collected. Whole saliva is stimulated by chewing on paraffin, and collected in a tube at the time-points of 0, 6, and 24 months. Parotid saliva is stimulated by 3% citric acid, and collected using Lashley’s cups at the start of the study and at 24 months.

Plaque bacteria samples are collected by rubbing foam pellets cervically on teeth 31-33, 34-35, and 36-37. This sampling is performed at three time-points: 0, 6, and 24 months.

In the prescreening, patients self-perform sampling at home with buccal swabs. Additionally, the dentist collects buccal swabs from the left and right buccal mucosa at baseline, 6 months, and 24 months.

Saliva, plaque samples, and swabs are kept at room temperature when collecting samples and data at the clinics. These samples are then interim stored at −20°C before they are transported by express delivery to the laboratory at Umeå University, for long-term storage at −80°C.

### REDCap collection of participant data

Participant data, including questionnaire and prescreen genotyping data, are collected and managed using the REDCap (Research Electronic Data Capture) tool hosted at Umeå University [31, 32]. The REDCap tool is a secure and intuitive web-based software and interface platform for clinical data capture, tracking, and management, including export procedures. This platform will allow investigators to access and follow the data from multiple clinics and laboratories.

The study questionnaire is sent to the participant as a digital link before their visit to the clinic, or can be completed at a visit to the clinic using a tablet. The questionnaire includes sociodemographic data (sex and ethnicity); and oral behaviour data, such as oral hygiene, intake frequency of sweets (e.g. cookies, biscuits, ice cream, or dried fruit) and sugary drinks (never, once per month, once per week, several times per week, once per day, several times per day), and the use of extra fluoride [6, 7].

### Data analysis and statistics

Questionnaire data will be downloaded directly from REDCap to excel files. Clinical outcome, genotyping, and other data will be entered into excel files, and then cleaned and secured in a database handled and stored at Umeå University by a professional data manager, with the same confidentiality as patient journal data. A head statistician at Umeå University will be responsible for data management and analysis. Data will be blinded in all analyses, as approved by the steering committee from detailed analyses plans.

### Data analyses plan for the basic Precaries-RCT

The primary outcome, ΔDeFS in the P4a^+^ arm, will be compared between the intensive and standard prevention groups, using the Mann‒Whitney U test with a significance level of 0.05. The primary intervention effect will be expressed as the % reduction in ΔDeFS caries increment. ΔDeFS = DeFS at the start of the study period subtracted from DeFS at the end of the study period, divided by the study period in years. A nonparametric test has been selected due to the expected positive skewness of data. The main analysis was performed according to the intent-to-treat principle. The secondary outcome, ΔDeFS in the P4a^−^ arm, will also use the Mann‒Whitney U test to test differences in ΔDeFS in P4a^−^ children. The Kruskal‒Wallis H test will be used to compare the ΔDeFS caries increment among the four groups. Pairwise post-hoc tests between groups will be performed using Dunn’s method. Socioeconomic and lifestyle-related variables will primarily be subject to descriptive statistical analysis.

### Health economic analyses

Health economic analyses will be performed to compare the total costs in each of the two arms of the trial, according to the levels of effectiveness for each arm. The incremental cost-effectiveness ratios (ICERs) will be calculated to provide an estimate of the mean cost per additional unit of outcome. Estimates of costs will also be included in the training and test samples for generation of cost-efficient healthcare using on-line multimodal machine learning.

### Precaries studies to support personalized dentistry

The Precaries trial includes the prospective RCT, adolescence [6, 7], and birth cohort [25] studies and samples (figures 1, 2 and 6, and supplemental table S2). Precaries-adolescence is a prospective case-referent sample of 452 adolescents followed from 12 to 17 years of age, for mining of human and microbiota target genes and pathways and omics data [6, 7]. The Precaries-birth cohort includes 650 families (trioses) that are being investigated by an interdisciplinary consortium for exposome and epigenetic factors programming health and caries, allergy, and neurophysiological development in childhood [25].

**Figure 6.**
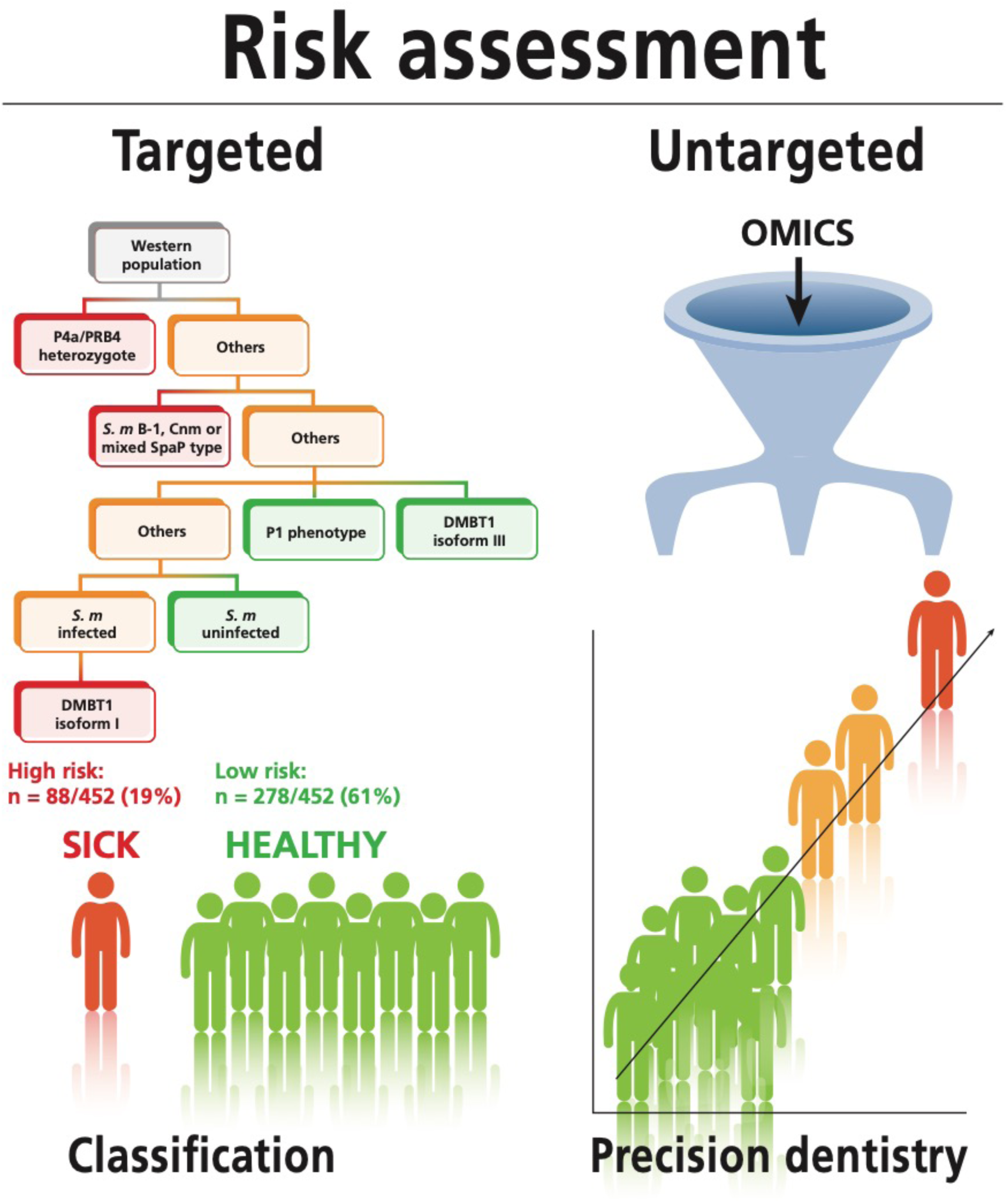
Targeted and untargeted strategies for risk assessment. Targeted risk assessment involves risk classification based on a set of causal human and microbiota genes and factors. Untargeted risk assessment involves precision dentistry based on omics data that allows for fine-tuned identification of the individual risk.

The Precaries-adolescence sample is a platform for targeted and untargeted multimodal omics approaches for risk classification and precision dentistry (figures 1 and 7, table 2). The targeted human predictor genes *PRH1-*2, *PRB1-4* and *DMBT1*, and *S. mutans spaP* and *cnm/cbm* genes, which are found to be useful for classification of healthy and sick individuals, will be further evaluated in the Precaries-RCT and Precaries-birth cohort.

Multimodal omics approaches—using data from proteomic, transcriptomic, glycogenomic, and genome-wide association studies (e.g. Global Screening Array–Multi-Disease) and human and microbiota genome sequences—will facilitate fine-tuned personalized assessment of risk and responsiveness to therapeutic agents. Both approaches will use quantitative imaging of the caries outcomes measures in the final models, and to generate refined clinical diagnoses.

Analyses will include whole-genome sequences and transcriptomes of *S. mutans* isolates (n=2500) and oral biofilm saliva and plaque samples, and saliva metabolomic, including short-chain fatty acid and mono- and disaccharide substrate, profiles. Potential therapeutics, including secondary metabolites, will be identified by filtering candidate gene structures against healthy and caries cases using bioinformatic tools [21, 22].

Targeted genes for metabolites and adhesin-receptor interactions, including SigLec and galectin glyco receptors, will be expressed or synthesized, and investigated for therapeutic potential [23, 24]. The ability of potential therapeutics to inhibit caries, inflammation and microbial clones will be examined using single-cell sequencing and the orthodontic caries model.

### Statistics and diagnostic platform

Analyses will comprise uni- and multivariate analysis and data mining, including clustering, hierarchical and machine learning models, depending on the data, outcome measures, and research questions. Nonparametric tests (e.g. Mann‒Whitney U and Kruskal‒Wallis H tests) will be used for groupwise comparisons related to caries outcomes at different time-points. Descriptive analyses will be initially performed to examine the distribution, rates, and saturation of outcomes. The ability of variables to explain and predict caries will be explored using mean/median difference with 95% CI, and fold-differences in prospective caries incidence and progression, and by using total multivariate partial least squares (PLS/OPLS) and other regression models. We will use the Precaries samples as training and test samples in cost-efficient healthcare approaches using multimodal machine learning.

### Patient and public involvement

It is well known to professionals and patients that prediction, treatment, and prevention are difficult among individuals with high caries activity, and that it is difficult to retain a caries-free status over time in some patients. The inability of biomarkers to predict caries risk, and the low efficacy of traditional prevention in the high-risk group are well-documented in systematic reviews by the Swedish Agency for Health Technology Assessment and Assessment of Social Services (SBU) [12, 14]. In terms of knowledge gaps, dentistry is the second largest category according to the SBU (Report to the Government, 2015-04-29), and this issue is addressed in the governmental investigation and report 2021 on “inequality of oral health” [#].

## DISCUSSION AND IMPACT

The basic Precaries-RCT will evaluate the efficacy of intensive and standard caries prevention, customized according to genotype and caries risk. The study will provide proof-of-concept for customized oral healthcare based on genetic profiling before lesions develop. Moreover, health economy analyses will indicate the cost-effectiveness of classifying individuals into healthy and sick trajectories, followed by customized prevention. The Precaries-RCT adaptive design, including self-performed sampling by mail, and the Precaries-adolescence platform will synergize in the development of cost-efficient oral healthcare using on-line multimodal machine learning.

The Precaries-adolescence sample is a platform for the mining of predictors and therapeutics using both targeted and untargeted omics approaches. It allows the identification of human genes and immunity pathways, and of different oro-types of the oral microbiota, which may be associated with genetic susceptibility/resistance and microbial dysbiosis and virulence/probiotic properties. Moreover, it allows hierarchical classification of omics data in machine and deep learning approaches, for fine-tuned individual risk prediction. Identifying underlying genes and molecules—including secondary metabolites, adhesins and receptors, and prebiotic molecules—will lead to the development of novel therapeutics, such as glycomimetics, that may influence the microbiota single-cell genomic profiles.

The Precaries-birth cohort allows implementation of diagnostic and therapeutic approaches in childhood, and with a trans-generational perspective. It also increases our understanding on how genetic and epigenetic factors synergize with the exposome in health and disease.

Finally, the orthodontic caries model applied in the Precaries-RCT allows for markedly shortened study times and measurements of caries induction, progression, and healing rates. Improved clinical criteria for healthy or caries-sick status is a vital part of precision dentistry.

### Ethics and dissemination

The present project is noninvasive, and conducted as part of the patient’s ordinary treatment at Public Dental Service clinics in several counties of Sweden. Treatment with orthodontic multibrackets is desired by the patients themselves for aesthetic and functional reasons, and the study inclusion and exclusion criteria follow the standard criteria at the clinics. Using a more sensitive caries scoring system (photo imaging and QLF) enables the identification of noncavitated caries lesions that are reversible, and will not harm the patient. All patients will be provided with basic fluoride exposure/prophylaxis, and if manifest caries occur, the treatment will be ended and the appliance will be removed. The study follows the generally accepted ethical principles of the Helsinki declaration, with regards to key values, such as life, health, dignity, integrity, right to self-determination, informed consent, privacy, confidentiality, dissemination of results, etc. The health and knowledge benefits outweigh any potential risks or burdens.

## Data Availability

All data produced in the present study are available upon reasonable request to the authors.

## Acknowledgments

The authors acknowledge the orthodontic assistants, clinicians, and all patients who participated in this study. We also acknowledge Sari Korva, Ola Wiodahl, Carina Öhman, and others for administrative support.

## Contributions

NS initiated and designed the study. NS, AW, and PL were responsible for the conception and design of the work. AW and NS drafted and finalized the manuscript, and all authors contributed to the manuscript and the study design, including the clinical, experimental, and analytical facets. All authors read and approved the manuscript before submission.

## Patient consent

Patient will be provided with written information, and will be asked to give their consent to participate in the study.

## Ethics approval

Dnr 2020-02533

## # Footnotes

Government investigation on equal oral health. When demand governs: An oral health system investigating equal oral health. Swedish government Office 2021. Available from: https://www.regeringen.se/rattsliga-dokument/statens-offentliga-utredningar/2021/03/sou-20218/

SBU [Knowledge and scientific knowledge gaps - Report to the Ministry of Health and Social Affairs]. Stockholm: Swedish Council on Health Technology Assessment in Health Care (SBU)]; 2015. Available from: http://www.sbu.se/contentassets/48f98e5ec9504a78af65b85bbb4c4e0e/kunskapsbehov-och-kunskapsluckor_sbu_s2014-8929-sam-delvis.pdf.

**Supplemental data**, separate file.

