## Supplemental files for "Personalized oral care (Precaries): an intervention study customized according to genetic cause and risk"

#### 1. Supplemental table S1

**Table 1. Time-points for enrolment, interventions and assessments in Precaries-RCT**

| | STUDY PERIOD | | | | |
|---|---|---|---|---|---|
| | Enrolment | Allocation | Post-allocation | | Close-out |
| **TIME-POINT** | -t1 | 0 | Base line | 6 m | 24 m |
| **ENROLMENT:** | | | | | |
| Eligibility screen | 2000 | | | | |
| Informed consent | 2000 | | | | |
| Human typing | 2000 | | | | |
| Lifestyle | 2000 | | | | |
| Baseline registration | 520 | | | | |
| Allocation | | 520 | | | |
| **INTERVENTIONS:** | | | | | |
| $P4a^{+}$, Intensive | | 65 | | | 50 |
| Standard | | 65 | | | 50 |
| $P4a^{-}$, Intensive | | 195 | | | 150 |
| Standard | | 195 | | | 150 |
| **ASSESSMENTS:** | | | | | |
| Lifestyle | | | X | X | X |
| Caries (DFS, DeFS) | | | X | X | X |
| Inflammation (BoP) | | | X | X | X |
| Microbiota/plaque | | | X | X | X |
| Microbiota/saliva | | | X | X | X |

### 2. Supplemental Table S2

**Table, 2. Outcome and measurements in the Precaries studies and samples**

| Precaries | Measurements/Samples | Time-points | | | | Outcomes | Comment | Purpose |
|---|---|---|---|---|---|---|---|---|
| **- 452 Adolescents** | | **12y** | | | **17y** | | | |
| Caries | Visual, tactile, X-ray | + | | | + | DeFS | Consensus caries index | ΔDeFS caries increment |
| Lifestyle | Questionairre | + | | | + | Oh, sweets | Oral hygiene (Oh) diet (sweets) | |
| Biological | Buccal swabs (DNA) | + | | | + | Human DNA | PRH1, PRH2 and DMBT1 genotying, glycogenomics, whole-genome sequencing, GSA-MD | Targets for predictors and therapeutics and for probiotic and dysbiotic microflora |
| | Parotid saliva | + | | | + | Transcriptome | | |
| | Whole saliva | + | | | + | Microbiome | 16S long read seq, metagenomics, single-cell metagenomics | |
| | Plaque | + | | | + | Microbiome | | |
| | *S. mutans* isolates | + | | | + | *S. m* population | Whole-genome seq & transcriptome | |
| **- RCT basic** | | **0** | | | **24m** | | | ΔDeFS and DeS increment, % caries reduction responders and non-responders to prevention |
| Caries | Visual, tactile, X-ray | + | | | + | DeFS, DeS | Buccal surface index (8 sites) | |
| | Photos | + | | | + | DeS | | |
| | QLF | + | | | + | DeS | | |
| Lifestyle | Questionairre | + | | | – | Oh, sweets | | |
| Biological | Buccal swabs (DNA) | + | | | – | $P4a^+$/$P4a^-$ | Self- and dentist swabings | |
| | Parotid saliva | + | | | – | | | |
| | Whole saliva | + | | | – | | | |
| | Plaque | + | | | – | | | |
| **- RCT adaptive** | | **0** | **6m** | **12m** | **24m** | | | |
| Caries | Visual, tactile, X-ray | + | + | + | + | DeS rate | Incidence and progression rates | see above |
| | Photos | + | + | + | + | DeS rate | | |
| | QLF | + | + | + | + | DeS rate | | |
| Lifestyle | Questionairre | + | – | – | + | Oh, sweets | | |
| Inflammation | Gingivitis | + | (+) | (+) | + | BoP, pocket formation | BoP = bleeding on probing periodontal pockets | Model for inflammation & periodontitis |
| Biological | Buccal swabs (DNA) | + | – | – | + | | | Evaluation and validation of human and microbiome genetic and transcriptome predictors |
| | Parotid saliva | + | – | – | + | | | |
| | Whole saliva | + | + | + | + | | | |
| | Plaque | + | + | + | + | | | |
| **- Birth cohort** | | **0** | **4m** | **1y** | **4y** | | | |
| Caries | Visual, tactile | – | – | – | + | DeFS | | |
| Exposome | Nutrients | + | + | + | + | | | Template for exposome and epigenetic factors and early programming and prevention during childhood |
| | Micronutrients | + | + | + | + | | | |
| | Toxic agents | + | + | + | + | | | |
| | Microbiome | + | + | + | + | | | |
| Biological | Swabs, saliva, plaque | – | – | – | + | | | |

#### 3. Participating clinics and clinicians, Precaries-RCT

- Umeå PhD Orofacial Medicine Lena Mårell
  Senior consultant in Orthodontics Charlotta Svanberg
  DDS Cariology Gustavo Silva
  Prosthodontic post graduate Erica Larsson
  Senior consultant in Cariology Maria André
  DDS Cariology Christina Radsjö Goriel

- Örnsköldsvik
  Senior consultant in Orthodontics Anna Erikson Lorenzo
  DDS Rakel Thrastardottir

- Gävle PhD, Senior consultant in Orthodontics Niels Ganzer
  PhD student, Senior consultant in Orthodontics Anke Krämer
  Orthodontic post graduate Caspar Carlfjord

- Örebro Associate Professor Orthodontics Farhan Bazargani
  Orthodontic post graduate student Samuel Andersson

- Linköping PhD, Senior consultant in Orthodontics Jenny Kallunki
  Orthodontic post graduate Mai Lin Lövgren

- Jönköping PhD Orthodontics Eva Josefsson
  Associate Professor Orthodontics Rune Lindsten
  PhD, Senior consultant in Orthodontics Anders Magnusson
  Orthodontic post graduate Isabell Hansson
  Orthodontic post graduate Erik Frilund

- Alingsås Senior consultant in Orthodontics Anna Tegnell
  DDS Haris Isic

- Mölndal Senior consultant in Orthodontics Victoria Granciuc
  Senior consultant in Orthodontics Elena Arezzo
  DDS Rebecka Akhlaghi

- Varberg Senior consultant in Orthodontics Seifi Esmaili
  Orthodontic post graduate Hanna Surac
  Orthodontic post graduate Reem Al-Taha

- Kristianstad Senior consultant in Orthodontics Brygida Gunwald
  Senior consultant in Orthodontics Henning Looström
  Orthodontic post graduate Firas Hittini
  DDS Nurije Kryeziu

Additional clinic´s

- Gothenburg Associate Professor Orthodontics Anna Westerlund
  PhD student Orthodontics Halah Khalifa
  DDS Ranna Yousif

- Malmö Associate Professor Orthodontics Mikael Sonesson

#### 4. Supplemental Figure S1

| Activity | Prevention | |
|---|---|---|
| | Intensified | Standard |
| Intervention time | 2 years | 2 years |
| Intervention period | 8 weeks | 8 weeks |
| Instruction – Follow-up | N=12 | N=12 |
| Information, instruction | 1 min at each recall N=12 | 1 min at each recall N=12 |
| Fluoride varnish (22 400 ppm) | N=6 | N=0 |
| Toothpaste two times daily | 5000 ppm | 1450 ppm |
| Renewal toothpaste | Every check-up | Every check-up |
| Renewal toothbrush | Every check-up | Every check-up |

**Supplemental Figure S1** Intensive and standard prevention given in the Precaries-RCT. The overall intervention time is 2 years, with 12 repeated prevention periods of 8 weeks each, including intensive prevention (IP) or standard prevention (SP). Each prevention period starts with instruction or reinstruction, and ends with a follow-up.

**5. Self-performed sampling of human DNA, Precaries-Screen (RCT)**

### Self-sampling

Swab of your cheek mucosa for caries research

#### Read through the entire instruction before sampling

This material is needed for the sampling:

---

Two test kits containing:

• Sampling swab

• Sample tube

Preparations before sample collection

---

For an instructional video of the sampling, scan the QR code with your mobile phone. Tap the box displayed on the screen to follow the link:
https://www.youtube.com/watch?v=Dh94gLJ4J5E.

Self-sampling

1. Take out the test kit containing the sample tube and swab. Open only one kit at a time.
2. Place the white end of the swab over the inside of one cheek, rotating it at least 10 times, or for about 30 seconds, pressing it firmly against the cheek's mucous membrane during sampling.
3. Insert the swab into the sample tube, with the white end down, and press the cap down to seal the tube. Pull out the green pin from the cap, and the white cushion will stick to the inside of the cap. Close the tube's lid with the plug that belongs to it, as shown in the image.
4. Repeat steps 1-3 with the inside of the other cheek.
5. Put the sample tubes in the reply envelope. Remove the adhesive strip and tightly seal the reply envelope. If the envelope is not adequately sealed, use tape to secure it.

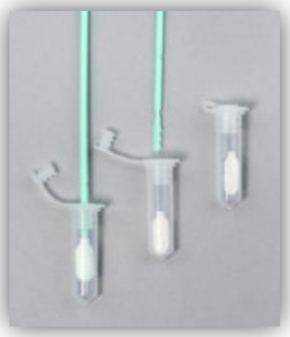

#### 6. Questionnaire Precaries-Screen (RCT)

Survey for the study "Evaluation of caries prevention based on genetic aetiology and risk"

**Instructions**
We would like you to answer a survey including questions about yourself. It is important that you answer the questions as accurately and honestly as possible. We want to emphasize that we, who work with the study, are all bound by confidentiality. Therefore, neither your school nor your parents will find out how you have answered these questions.
Unless otherwise indicated, put a checkmark in one of the boxes. If there is anything you wonder about or do not understand, ask someone on the staff at the dental clinic.
Thank you for your participation!

**Questions about yourself**

1. Are you a
   - Boy ☐
   - Girl ☐
   - Other ☐

2. Were you born in a country other than Sweden?
   - ☐ No
   - ☐ Yes, which country? ........................................

3. Do you have a parent who has immigrated to Sweden?
   - ☐ Yes, both, from which country?.................................................................
   - ☐ Yes, one, from which country? ……………………………………………
   - ☐ Yes, several (e.g. step-parents)
   - ☐ No, non

4. You live in Sweden. We wonder if you feel Swedish or if you feel like something else—Roma, Turkish, Sami, Iranian, Finnish, etc.
   - ☐ Swedish
   - ☐ Other, what?

5. Where do you live?
   - ☐ Big city or suburb of a big city
   - ☐ City
   - ☐ Larger community
   - ☐ Village/rural area

Dental health questions

10. How often do you brush your teeth?
    - ☐ More than twice a day
    - ☐ Twice a day
    - ☐ Once a day
    - ☐ Less than once a day/irregularly

11. Do you use toothpaste?
 Yes
 No

12. Is it fluoride toothpaste?
 Yes
 No
 Don't know

13. Do you use a regular toothbrush or an electric toothbrush?
 Regular
 Electric
 Switch between regular and electric

14. Do you use anything other than a toothbrush to clean your mouth? (You can check several options!)
 No
 Yes, dental floss occasionally
 Yes, dental floss almost every day (about 4 times a week)
 Yes, mouthwash

15. Do you usually use fluoride in products other than toothpaste? (You can check several options!)
 No
 Yes, I rinse with fluoride solution more than once a month
 Yes, I chew fluoride gum, 1-3 pieces/day
 Yes, I take fluoride tablets, 1-3 tablets/day

**Questions about diet**

16. Cakes, pastries, ice cream, candy, and dried fruit are tasty; how often do you eat any of these things?
 Never
 Once a month
 Once a week
 Several times a week
 Once a day
 Several times a day

17. How often do you drink juice, sports drinks, or soda?
 Never
 Once a month
 Once a week
 Several times a week
 Once a day
 Several times a day

18. How much do you weigh? .....................kg

19. How tall are you? .....................cm

#### 7. Prevention, intensified and standard, clinic protocol

### Intensive prevention group

- Duraphat toothpaste and toothbrush are provided at the appointment
- Oral hygiene instructions according to routine
- 3 photos edge-to-edge at each visit, every 8 weeks
- D-lack every other visit, i.e. every 12 weeks
- New toothpaste approximately every 6 weeks
- New toothbrush approximately every 3 months
- Bleeding on probing index in the 3rd quadrant m, b, d

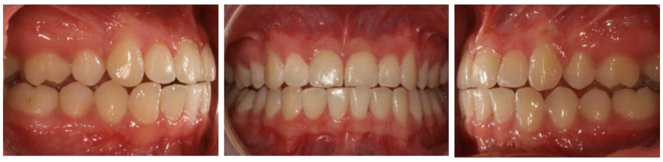

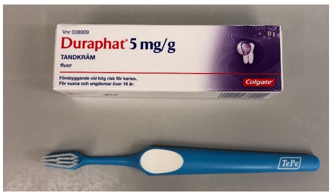

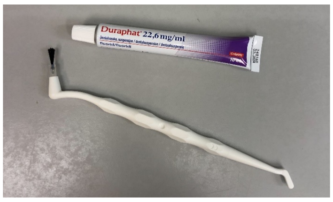

### Standard prevention group

- Colgate toothpaste and toothbrush are provided at the appointment
- Oral hygiene instructions according to routine
- 3 photos edge-to-edge at each visit, every 8 weeks
- New toothpaste approximately every 6 weeks
- New toothbrush approximately every 3 months
- Bleeding on probing index in the 3rd quadrant m, b, d

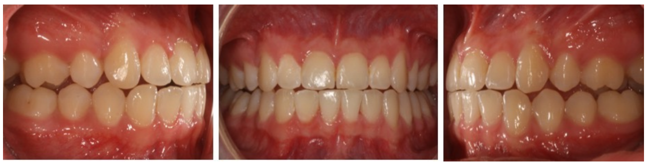

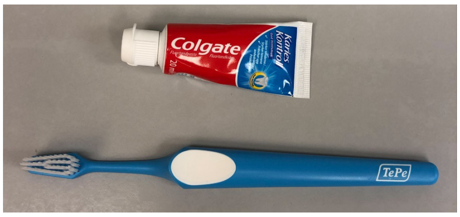

#### 8. Compliance questionnaire, Precaries-RCT

1. How often do you brush your teeth?
   ☐ More than twice a day
   ☐ Twice a day
   ☐ Once a day
   ☐ Less than once a day/irregularly

2. Do you use a regular toothbrush or an electric toothbrush?
   ☐ Regular
   ☐ Electric
   ☐ Switch between regular and electric toothbrush

3. Do you use anything other than a toothbrush to clean your mouth? (You can check multiple options!)
   ☐ No
   ☐ Yes, dental floss occasionally
   ☐ Yes, dental floss almost every day (about 4 times a week)
   ☐ Yes, mouthwash

4. Do you use toothpaste?
   ☐ Yes
   ☐ No

5. Do you use the toothpaste that you received from the clinic?
   ☐ Yes
   ☐ No
   ☐ Don't know

6. How often do you use the toothpaste that you received from the clinic?
   ☐ More than twice a day
   ☐ Twice a day
   ☐ Once a day
   ☐ Less than once a day/irregularly

7. Do you use fluoride in any other way than toothpaste? (You can check multiple options!)
   ☐ No
   ☐ Yes, I rinse with fluoride solution more than once a month
   ☐ Yes, I chew fluoride gum, 1-3 pieces per day
   ☐ Yes, I take fluoride tablets, 1-3 tablets per day.

**9. Caries calibration, Precaries-RCT, dentist protocol**

### Caries calibration

Program for calibration of dentists for caries research
The program comprises two parts: radiological and clinically patient-centred calibration.

#### Radiological

- Powerpoint 1: Inter-individual calibration. Cases of 10 patients with X-ray bitewing
- Powerpoint 2: Inter-individual calibration. Cases of 10 patients with X-ray bitewing
- Powerpoint 3. Inter-individual calibration. Cases of 10 patients with X-ray bitewing
- Powerpoint 4: Same as the first step "Powerpoint 1", to assess intra-individual assessment and variation
- Note the registrations in the attached templates and compare between participating dentists

#### Clinical patient-centred

- Schedule child patients for revision examination (n=8-12). For the PreCaries project, the patients' ages should be between 14-25 years.
- Note the registrations in the attached templates, and compare them between participating dentists, as a reference.

#### Evaluation

- Kappa values and percent agreement is calculated.

Use the following protocol

Fall N: Fall N:

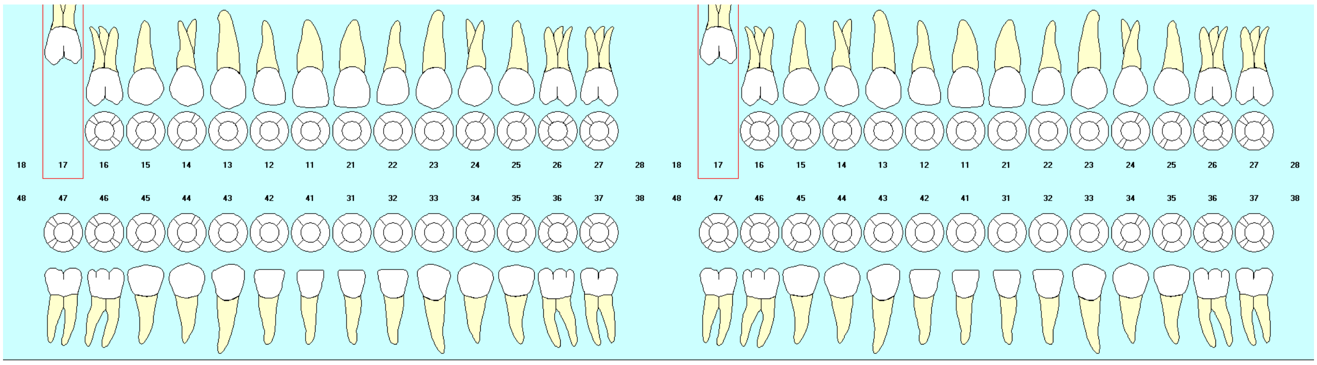

### 10. DeFS-registration, Precaries - RCT

Name/ID: ______________________________________________

Date: ________________________________________________

Caries (0-3)
Fillings (F)
Mineralization disturbances (M)

#### Caries, mineralization disturbances, fillings

**DeFS-index**

| | | 17 | 16 | 15 | 14 | 13 | 12 | 11 | 21 | 22 | 23 | 24 | 25 | 26 | 27 | |
|---|---|---|---|---|---|---|---|---|---|---|---|---|---|---|---|---|
| | **m** | | | | | | | | | | | | | | | **m** |
| | **o** | | | | | | | | | | | | | | | **o** |
| | **d** | | | | | | | | | | | | | | | **d** |
| | **l** | | | | | | | | | | | | | | | **l** |
| | **b** | | | | | | | | | | | | | | | **b** |
| | **Buccal surface (8-surface index)** | | | | | | | | | | | | | | | |
| B | d | | | | | | | | | | | | | | | d |
| U | bd | | | | | | | | | | | | | | | bd |
| C | c | | | | | | | | | | | | | | | c |
| C | bc | | | | | | | | | | | | | | | bc |
| A | m | | | | | | | | | | | | | | | m |
| L | bm | | | | | | | | | | | | | | | bm |
| | i | | | | | | | | | | | | | | | i |
| | bi | | | | | | | | | | | | | | | bi |

**DeFS**

| | | 47 | 46 | 45 | 44 | 43 | 42 | 41 | 31 | 32 | 33 | 34 | 35 | 36 | 37 | |
|---|---|---|---|---|---|---|---|---|---|---|---|---|---|---|---|---|
| | **m** | | | | | | | | | | | | | | | **m** |
| | **o** | | | | | | | | | | | | | | | **o** |
| | **d** | | | | | | | | | | | | | | | **d** |
| | **l** | | | | | | | | | | | | | | | **l** |
| | **b** | | | | | | | | | | | | | | | **b** |
| | **Buccal surface (8-surface index)** | | | | | | | | | | | | | | | |
| B | d | | | | | | | | | | | | | | | d |
| U | bd | | | | | | | | | | | | | | | bd |
| C | c | | | | | | | | | | | | | | | c |
| C | bc | | | | | | | | | | | | | | | bc |
| A | m | | | | | | | | | | | | | | | m |
| L | bm | | | | | | | | | | | | | | | bm |
| | i | | | | | | | | | | | | | | | i |
| | bi | | | | | | | | | | | | | | | bi |

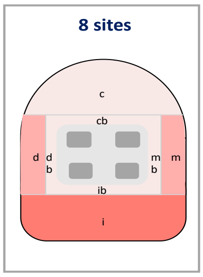

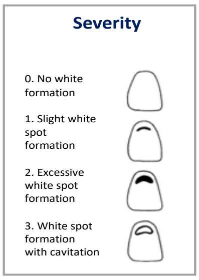

DeFS: ________________________________

8 surfaces:_____________________________

Mineralization disturbances: _________________

Name/ID: ___________________________________________

Date: _____________________________________________

**BoP**- Put a cross where it's bleeding and mark "99" for extracted teeth

| | 31 | 32 | 33 | 34 | 35 | 36 | 37 |
|---|---|---|---|---|---|---|---|
| m | | | | | | | |
| b | | | | | | | |
| d | | | | | | | |

BoP (n):_________________

BoP (%): _________________

| Whole Saliva (HS) | | |
|---|---|---|
| Volume (mL) | Time (min.) | Secr. (mL/min) |

#### 11. Procedure for baseline examination, Precaries-RCT

1. **Clinical sampling**
   A. Buccal swab
   The inside of each cheek is swabbed with a rotating swab. Each cheek is swabbed at least 10 times with a separate swab. The swabs are air-dried for 15 minutes, and then stored in their original packaging, labelled with a sticker.
   B. Plaque sampling (PL)
   Dry-blast quadrant 3. Scrape plaque from the buccal surfaces of teeth 31-33, 34-35, and 36-37 with a sterile foam pellet (PL1, PL2, and PL3, respectively). Transfer the foam pellets to a 2-mL tube with sterile 15% glycerol and glass beads.
   C. Chewing-stimulated whole saliva
   The patient sits slightly forward and chews on a paraffin pellet until the pellet becomes soft, and then the patient is asked to swallow all saliva. The patient is then asked to start chewing. Timing starts when the patient spits for the first time. All produced saliva is continuously collected through a funnel into a test tube until 3 mL is obtained, and the time is noted.
   D. Parotid saliva (PS)
   Place the patient in a semi-reclined position, and locate the openings of the parotid ducts. Connect the Lashley cups to the suction device, and ensure that there is suction in both cups. Apply the cups over the parotid ducts. Have the patient hold the suction tubing to keep the cups in place, and encourage them not to clench their cheeks. Begin stimulation by rubbing 3% citric acid over the tongue. Then place their openings in a 15-mL Falcon tube, and start timing when saliva begins to flow out. Stimulation is repeated until 3 mL is obtained, and the time is noted.
2. Registration of Bleeding on Probing index (BoP) at teeth 31-33, 34-35, and 36-37.
3. 4 bitewing X-rays
4. Polishing if necessary
5. Registration of caries DeFS, 8-surface index, and mineralization disorders. See protocol
6. Clinical photographs; 5 intraoral photos
7. QLF photography; 5 intraoral photos
8. Scan and store the protocol in Sharepoint in the "Caries" and "BoP" folders for the respective clinic and time. Label the folder with ID and name.
9. Mark in REDCAP that the baseline check-up has been done.
10. Edit photos and insert them into the respective folder and PowerPoint.

**Sample handling after collection**

• Samples should be placed in the freezer as soon as possible.
• Mark with a thin water-resistant pen in a legible manner, ensuring that it does not smear.

A. Plaque sampling (PL1-3). The sample tubes are vortexed for at least 1 minute, and it is ensured that the plaque is broken down by glass beads to obtain a homogeneous solution. Transfer 250 µL of the solution to a new tube using a pipette. Both tubes are labelled and stored in a −20°C freezer at the clinic.
B. Saliva is vortexed for approximately 5 s, and then aliquoted into 2-mL sterile tubes (1 mL per tube). All test tubes are labelled and stored in a −20°C freezer at the clinic.
C. Parotid Saliva (PS). Saliva is vortexed for about 5 seconds, and then aliquoted into 2-mL sterile tubes with hooks (1 mL per tube). All tubes are labelled and then stored in a −20°C freezer at the clinic.
D. The Lashley cups are washed with hand dishwashing detergent, the hoses are rinsed with tap water, and then packed and autoclaved overnight.
E. The funnels are washed with hand dishwashing detergent and air-dried overnight.

#### 12. Procedure for 6-month examination, Precaries-RCT

1. **Clinical sampling**

A. Buccal swab
The inside of each cheek is swabbed with a rotating swab. Each cheek is swabbed at least 10 times with a separate swab. The swabs are air-dried for 15 minutes, and then stored in their original packaging, labelled with a sticker.

B. Plaque sampling
Dry-blast quadrant 3. Scrape smear from the buccal surfaces of teeth 31-33, 34-35, and 36-37 with a sterile foam pellet (PL1, PL2, and PL3, respectively). Transfer the foam pellets to a 2-mL tube with sterile 15% glycerol and glass beads.

C. Chewing-stimulated whole saliva
The patient sits slightly forward and chews on a paraffin pellet until the pellet becomes soft, and then the patient is asked to swallow all saliva. The patient is asked to start chewing. Timing starts when the patient spits for the first time. All produced saliva is continuously collected through a funnel into a test tube until 3 mL is obtained, and the time is noted.

2. Registration of Bleeding on Probing index (BoP) at teeth 31-33, 34-35, and 36-37.
3. Removal of apparatus (everything except bracket)
4. 4 bitewing X-rays
5. Polishing if necessary
6. Registration of caries DeFS, 8-surface index, and mineralization disorders. See protocol
7. Clinical photographs; 5 intraoral photos
8. QLF photography; 5 intraoral photos
9. Compliance questionnaire to be completed
10. Mark in REDCAP that the 6-month check-up has been done
11. Scan and store the protocol in Sharepoint in the "Caries" and "BoP" folders for the respective clinic and time. Label the folder with ID and name.
12. Edit photos and insert them into the respective folder and PowerPoint.

**Sample handling after collection**

• Samples should be placed in the freezer as soon as possible.
• Mark with a thin water-resistant pen in a legible manner, ensuring that it does not smear.

A. Plaque sampling (PL1-3). The sample tubes are vortexed for at least 1 minute, and it is ensured that the plaque is broken down by glass beads to obtain a homogeneous solution. Transfer 250 µL of the solution to a new tube using a pipette. Both tubes are labelled, and stored in a −20°C freezer at the clinic.

B. Chewing-stimulated whole saliva (HS). Saliva is vortexed for approximately 5 s, and then aliquoted into 2-mL sterile tubes (1 mL per tube). All test tubes are labelled and stored in a −20°C freezer at the clinic.

C. The funnels are washed with hand dishwashing detergent, and air-dried overnight.

#### 13. Procedure for 24-month examination (debond), Precaries-RCT

1. **Clinical sampling**
   A. Buccal swab
   The inside of each cheek is swabbed with a rotating swab. Each cheek is swabbed at least 10 times with a separate swab. The swabs are air-dried for 15 minutes and then stored in their original packaging, labelled with a sticker.
   B. Plaque sampling
   Dry-blast quadrant 3. Scrape smear from the buccal surfaces of teeth 31-33, 34-35, and 36-37 with a sterile foam pellet (PL1, PL2, and PL3, respectively). Transfer the foam pellets to a 2-mL tube with sterile 15% glycerol and glass beads.
   C. Chewing-stimulated whole saliva
   The patient sits slightly forward and chews on a paraffin pellet until the pellet becomes soft, and then the patient is asked to swallow all saliva. The patient is asked to start chewing. Timing starts when the patient spits for the first time. All produced saliva is continuously collected through a funnel into a test tube until 3 mL is obtained, and the time is noted.
   D. Parotid saliva (PS)
   Place the patient in a semi-reclined position, and locate the openings of the parotid ducts. Connect the Lashley cups to the suction device, and ensure that there is suction in both cups. Apply the cups over the parotid ducts. Have the patient hold the suction tubing to keep the cups in place, and encourage them not to clench their cheeks. Begin stimulation by rubbing 3% citric acid over the tongue. Then place their openings in a 15-mL Falcon tube, and start timing when saliva begins to flow out. Stimulation is repeated until 3 mL is obtained, and the time is noted.
2. Registration of Bleeding on Probing index (BoP) at teeth 31-33, 34-35, and 36-37.
3. Removal of appliance
4. 4 bitewing X-rays
5. Polishing if necessary
6. Registration of caries DeFS, 8-surface index, and mineralization disorders. See protocol
7. Clinical photographs; 5 intraoral photos
8. QLF photography; 5 intraoral photos
9. Mark in REDCAP that the debond examination has been done.
10. Scan and store the protocol in Sharepoint in the "Caries" and "BoP" folders for the respective clinic and time. Label the folder with ID and name.
11. Edit photos and insert them into the respective folder and PowerPoint.

**Sample handling after collection**

• Samples should be placed in the freezer as soon as possible.
• Mark with a thin water-resistant pen in a legible manner, ensuring that it does not smear.

A. Plaque sampling (PL1-3) The sample tubes are vortexed for at least 1 minute, and it is ensured that the plaque is broken down by glass beads to obtain a homogeneous solution. Transfer 250 µL of the solution to a new tube using a pipette. Both tubes are labelled and stored in a −20°C freezer at the clinic.
B. Saliva is vortexed for approximately 5 s, and then aliquoted into 2-mL sterile tubes (1 mL per tube). All test tubes are labelled and stored in a −20°C freezer at the clinic.
C. Parotid saliva (PS). Saliva is vortexed for about 5 seconds and then aliquoted into 2-mL sterile tubes with hooks (1 mL per tube). All tubes are labelled and then stored in a −20°C freezer at the clinic.
D. The Lashley cups are washed with hand dishwashing detergent, the hoses are rinsed with tap water, and then packed and autoclaved overnight.
E. The funnels are washed with hand dishwashing detergent, and air-dried overnight.
